# Single Site performance of AI software for stroke detection and Triage

**DOI:** 10.1101/2021.04.02.21253083

**Authors:** Dan Paz, Daniel Yagoda, Theodore Wein

## Abstract

**Background:** Recently developed software utilizing artificial intelligence for fast detection and triage of stroke cases has the potential to accelerate stroke care and improve patient outcomes. We performed this analysis to evaluate the performance and time-to-notification of one such software - RAPID LVO.

**Methods:** We created a database of 151 consecutive acute stroke patients for whom CT scans were processed by the RAPID LVO software over a period of eight months. The LVO notification and time to notification of the software were collected, alongside patient information and the CTA findings.

**Results:** RAPID LVO achieved a sensitivity of 63.6% and specificity of 85.8% for large vessel occlusion, with an average time to notification of 32.53 minutes.

**Conclusions:** RAPID LVO has low sensitivity, moderate specificity and high time-to-notification performance. Our study data demonstrated in particular low overall sensitivity (63%) for distal occlusions (M2-3). The disparity between the observed performance and the performance reported in RAPID LVO’s FDA clearance demonstrates the importance of independent, multi-center evaluation. The gap between the performance in this study compared to published records of RAPID AI may be due to differences in imaging hardware, software implementation, connectivity or clinical definitions.

## Introduction

Stroke is the leading cause of serious disability and death worldwide, with an annual incidence exceeding 15 million and a prevalence of 42 million ^1^. In the USA alone, stroke is affecting nearly 800,000 people annually, with up to 3% of the US population reported they were disabled because of stroke^2,3^.

Large vessel occlusions (LVOs) are defined as acute occlusion of the proximal intracranial anterior and posterior circulation, causing hypoperfusion in the affected cerebral territory, which is presented clinically as an acute stroke. The vast majority of strokes are ischemic strokes, and LVOs account for 24-46% of them^4^.

Effective therapies such as intra-arterial thrombolysis or mechanical thrombectomy are available for patients suffering from LVOs. However, The efficacy of these treatments is highly time sensitive, as was demonstrated in numerous randomized controlled trials and meta-analysis studies^5,6,7,8,9,10^, with a single minute of delay translating to the loss of 1.9M neurons^11^, 4.2 days of disability^12^and $1,059 of lost net economic benefit to the health system^13^. Hence, timely diagnosis of a stroke patient suffering from LVO is crucial for treatment and for a better prognosis.

Recent advances in artificial intelligence, cloud computing and mobile technology enabled the development of novel medical software that automatically analyzes vascular imaging studies and alerts entire care teams simultaneously on suspected LVOs.

Automatic detection is especially crucial in medical institutions which do not have neuroradiologists available 24 hours a day and 7 days per week.^14^. These sites typically use overnight coverage by general radiologists (either in-house or via Teleradiology services), who are less specialized than neuroradiologists in the interpretation of advanced neurovascular imaging and the detection of a LVO stroke, resulting in lower accuracy and longer time to detection. Automatic triage systems are intended to bridge this gap and provide timely notifications directly to the neurovascular specialist on-call.

In recent months, several publications demonstrated the real-world impact of one such system, Viz LVO (Figure 1, left), demonstrated a reduction in time-to-treatment, reduced length of stay and improved patient outcomes, when comparing pre- and post-implementation of the AI software. In one study, Hassan et al.^15^ have found a significant reduction of 66 minutes in the median time from CTA acquisition at a primary stroke center (PSC) to arrival at a comprehensive stroke center (CSC), and a reduction of 2.5 days in the length of Neuro-ICU stay (p=0.00086) when comparing pre- and post-Viz LVO implementation. Morey et al.^16^ demonstrated both statistically significant time savings as well as statistically significant improvement in patient outcomes, as measured by NIHSS at discharge as well as 5-day and 90-day follow-ups. These increased efficiencies, and their impact on patient outcomes, are in line with current understanding of the complexities and bottlenecks of stroke workflow^17^.

**Figure 1:**
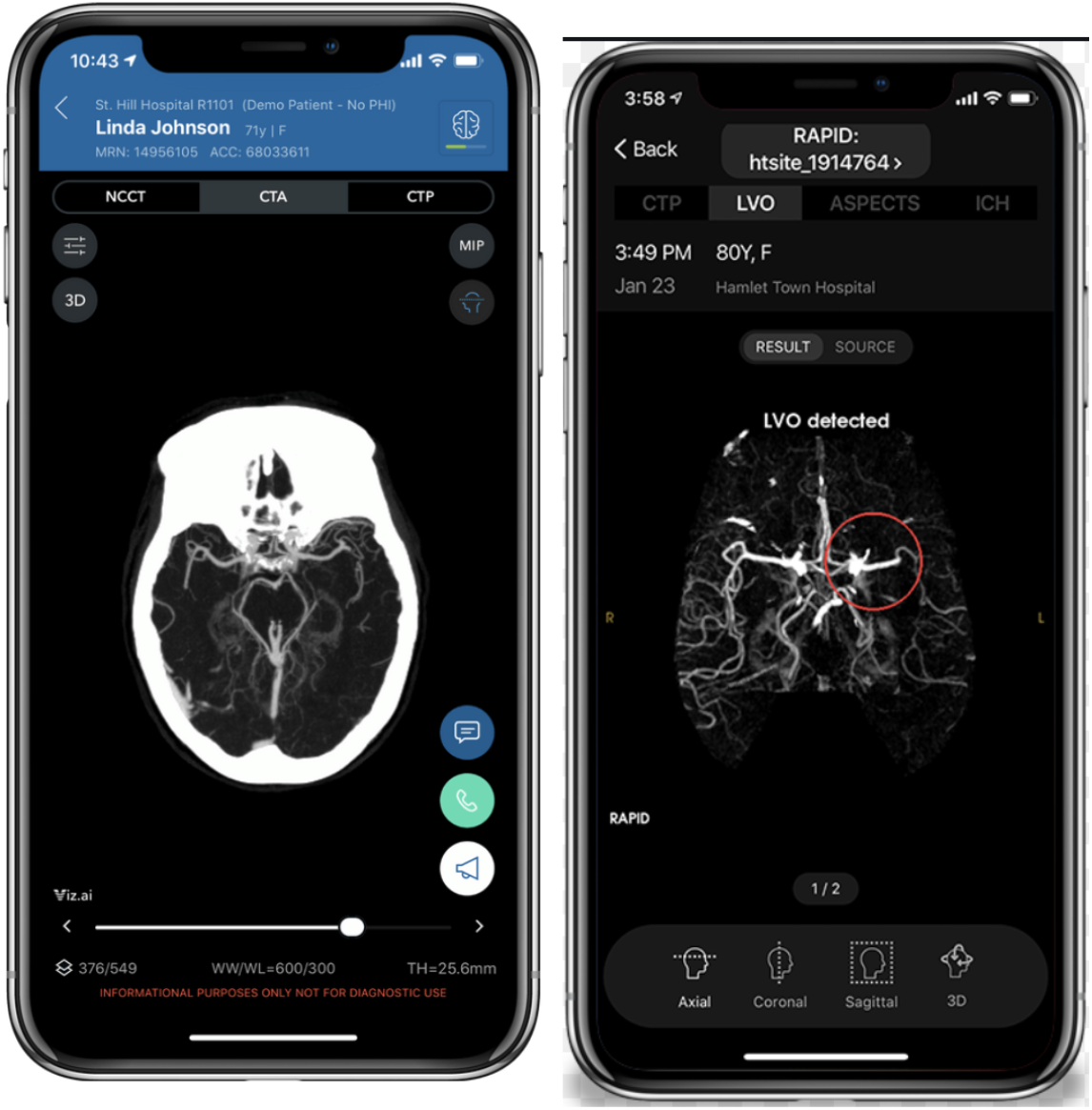
software for automatic detection and triage of large vessel occlusions from CTA. **left:** Viz.ai application, **right:** RAPID.AI application.

Recently, a new LVO detection software was launched commercially by RAPID.AI, called RAPID LVO (Figure 1, right). We have been using RAPID LVO for several months, and the purpose of this study is to evaluate the performance of the software in terms of sensitivity, specificity and processing time to establish its clinical utility. To the best of our knowledge, this is the first independent evaluation of the performance of RAPID LVO.

## Methods

A retrospective study was conducted including patients who presented with suspected acute stroke symptoms at the Montreal neurological hospital (MNH). All patients whose imaging studies were processed by RAPID LVO between the dates 4/7/20 and 31/12/20 were included in the study. The AI Software utilized in this study was RAPID LVO by iSchemiaview Inc. The software analyzes CT angiogram images of the brain, acquired in the acute setting, and sends notifications that a suspected large vessel occlusion has been identified. Images can then be accessed through the mobile application.

The study was approved by the local Institutional Review Board-MUHC REB NEUPSY.

### Imaging protocol

All the patients included in the study underwent imaging, and subsequently, thrombectomy procedure if needed, at the same tertiary stroke center. The institutional stroke protocol, which was performed in all patients in the study, consists of non-contrast agent enhanced CT acquisition of the head, followed by acquisition of contrast enhanced CT (section thickness, 0.8mm-1mm) of the head. The basic imaging used 80kv and varying mA values for the 19 volumes acquired: 310mA for the mask, 150mA during pre-arterial phase, 300mA during arterial phase and finally 150mA for the rest of the acquisition. Rotation time was 0.75. The CT scanner that was used is the Toshiba Aquilion One 320 slices. The contrast that was used was ISOVUE 370. Total scan time was 60 seconds. The servers were the Internal PACS and RAPID server. Reformats were MPR on arterial peak. Axial thickness 1mm, with interval of 0.8mm. MIP on all 19 volumes coronal and sagittal 2mm, with 2mm interval, DSA movie and Perfusion maps.

### Data and statistical analysis

Demographic and clinical variables as well as the software’s output and processing times were collected. Demographic variables include date, age and sex; clinical variables include the presence or absence of a LVO as well as its location and the affected side.

The statistical analysis was carried out as follows. For quantitative variables, averages and standard deviations are reported. For binary or categorical variables, percentages are reported. Each patient is defined as either a true positive, true negative, false positive or a false negative based on the clinical variables and the output of the software (see Table 2 for more details). Two cases where the software alerted, but identified the occlusion on the wrong side, were counted as False Positives.

**Table 2:** Definitions of terms used for correlating ground truth and software output

|  | LVO YES | LVO NO |
| --- | --- | --- |
| RapidAI alert<br>YES | True Positive | False Positive |
| RapidAI alert<br>NO | False Negative | True Negative |

Sensitivity and specificity are estimated as proportions, and 95% two-sided confidence intervals are computed using the Clopper-Pearson method. Time-to-notification was defined as the duration of time from the start of the scan (defined as the first scout) to arrival of the notification. The average is reported as well as a 95% confidence interval computed using the t distribution. All computations were performed using Google Sheets and the programming language Python3.7.

## Results

A total of 151 patients were collected during the study (mean age 70.57 (±15.87), 54.3% women). Of these patients, 85 did not have LVOs and 66 did have LVOs (51.5% on left side, 45.4% right side, 3% posterior circulation). LVO locations were as follows: ICA (4 patients), MCA-M1 (32 patients), MCA-M2/3 (28 patients), posterior circulation (2 patients). A single Tandem ICA-M1 occlusion was counted as MCA-M1. A total of 54 alerts were generated by the software, with 42 being true positive alerts and 12 being false positive alerts. Examples are provided in Figure 2.

**Figure 2:**
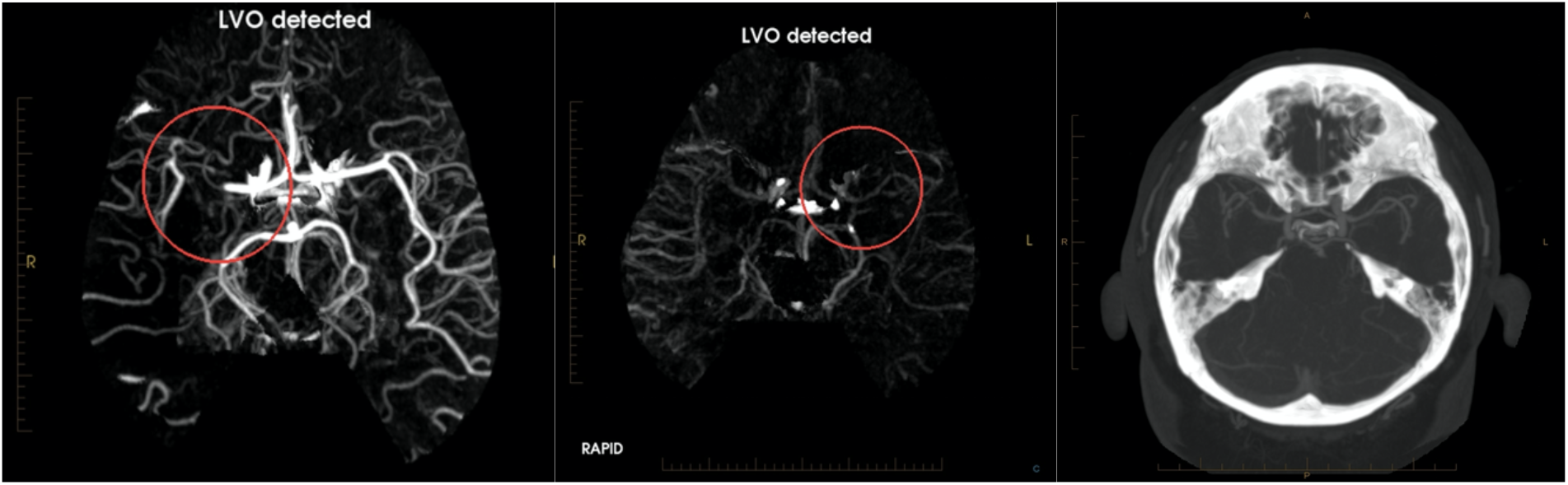
examples of true positive and false positive alerts. Left: A true positive alert, detecting a right MCA-m1 occlusion. Middle: False positive alert, detecting a non-existing left MCA-M1 occlusion. Right: Maximal intensity projection (MIP) of the original CTA for which the false positive alert was generated, demonstrating potent left MCA-M1.

Overall sensitivity and specificity were 62.12% [49.34, 73.78] and 85.88% [76.64, 92.49], respectively. In two cases, the software generated an alert, but the visual marker indicated the wrong location (see Figure 2, middle). These cases were counted as false-positives. Average time to notification was 32.53 minutes (sd. 63.11 mins). These results are summarized in Table 1. There were 3 cases whose alerts had arrived in a delay of a few hours or days. When excluding these patients, the average time to notification was 18.31 minutes (sd. 5.07mins).

**Table 1:**
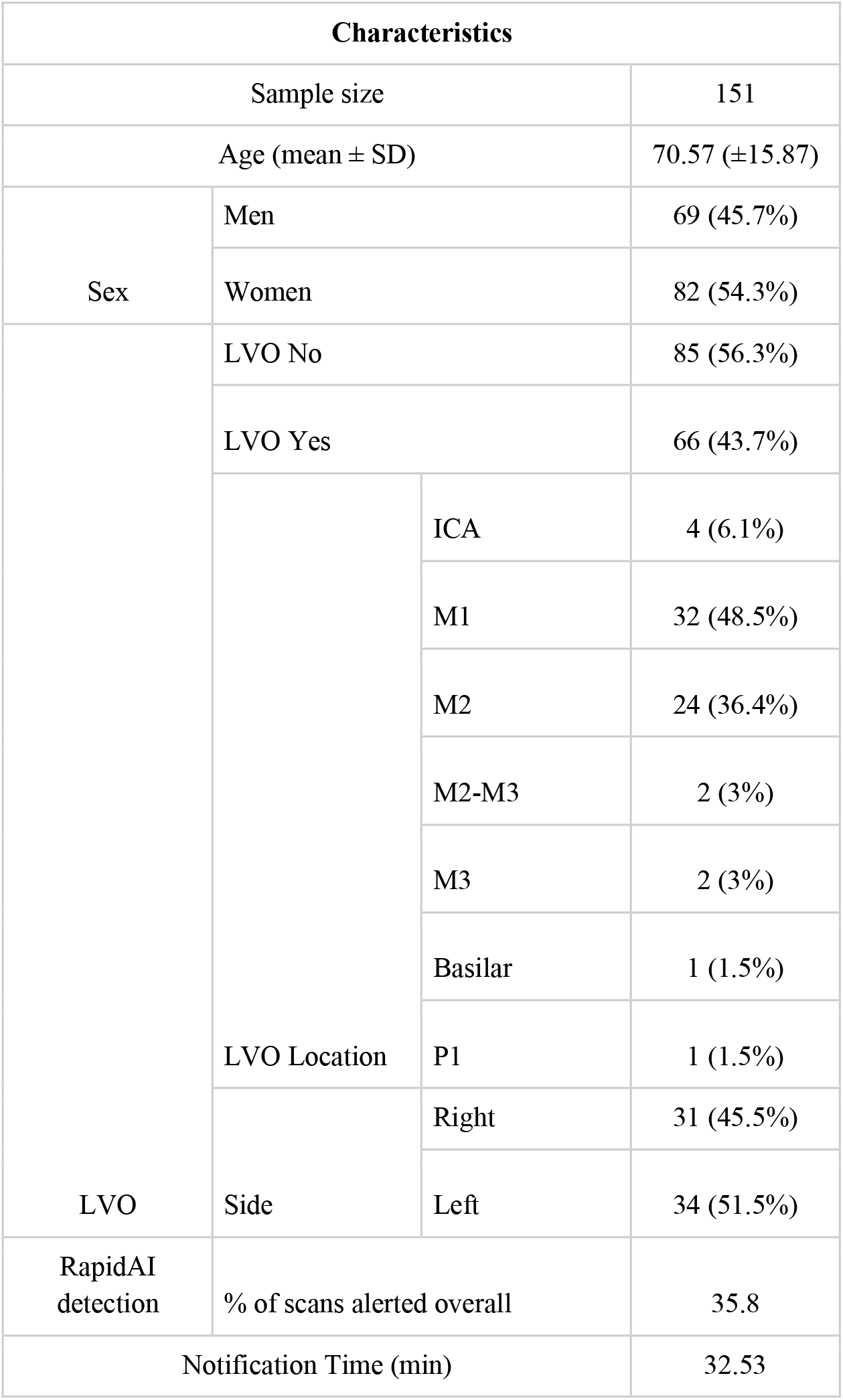
summary of the variables collected in this study

Remarkably, the estimated performance metrics are different from other publications describing the performance of the device. For example, the performance of the device as reported in the relevant FDA clearance^18^ or by Amukotuwa et al.^19^. These different estimates are summarized in Table 2.

**Table 2:** comparison of the performance of RAPID LVO as published in the relevant regulatory clearance vs. the performance observed in this study.

| Performance metric | RAPID LVO FDA clearance | Amukotuwa et al. | Our study |
| --- | --- | --- | --- |
| Sensitivity (%) | 0.970 [0.933,0.987] | 0.969 [0.943,0.985] | 0.636 [0.508, 0.751] |
| Specificity (%) | 0.956 [0.919, 0.977] | 0.743 [0.706,0.778] | 0.858 [0.766, 0.924] |
| Time-to-Notification (min) | 2.86 [2.79, 2.92] | NA | 18.3 (sd. 63.11) |

Among the 12 false positive results, except for one study all of the CTA were considered technically good. In two studies a subdural hematomas were detected, a unilateral in one study and bilateral on the other study. In two out of the false positive cases no possible cause for the false detection was found. In the remaining of the false positive cases different degrees of stenosis were found either in the ICA or M1 stenosis. The findings are summarized in Table 3.

**Table 3:** Summary of the false positive cases and possible findings that influence the end result

| No | Technical quality of the study | Possible finding that can influence the end result | Remarks |
| --- | --- | --- | --- |
| 1 | Good | Bilateral SDH |  |
| 2 | Good | Non | 25% ICA stenosis |
| 3 | Good | Severe Distal ICA stenosis |  |
| 4 | Good | Severe proximal ICA stenosis |  |
| 5 | Moderate motion artifact | Chronic ICA occlusion |  |
| 6 | Good | Acute on chronic ICA occlusion |  |
| 7 | Good | Severe distal M1 stenosis |  |
| 8 | Good | Non |  |
| 9 | Good | Unilateral SDH |  |
| 10 | Good | Severe P2 stenosis |  |
| 11 | Good | Severe M1 stenosis |  |
| 12 | Good | Severe M1 stenosis | LVO was noted on the contralateral side |

## Discussion

This study evaluated the RAPID AI software which analyses CTA scans of suspected stroke patients to provide an automatic alert of an intracranial circulation LVOs. The software demonstrated a sensitivity of 63.6% and specificity of 85.8%, as well as time-to-notification of over 18.3 minutes, on average. There is a remarkable gap between the performance witnessed in this study and recent publications Amukotuwa et al ^19^, including the formal numbers provided in the software developers. This gap, coupled with the long time-to-alert (18.3 minutes, on average) may reduce the clinical efficacy of the device in the emergent clinical where fast and accurate decision making is a key factor for a better prognosis.

In comparison, Viz LVO is a different software Shalitin et al. ^20^evaluated the performance of Viz LVO by aggregating data from 139 sites, and had found overall sensitivity and specificity of 96% and 93%, respectively, and an average time to notification of 5:45 minutes.

Lev et al.^21^estimated the sensitivity and specificity of neuroradiologists reading scans for LVOs at 98% and 86%, respectively, and estimated the time-to-analysis at 15 minutes. More recently, Becks et al.^22^ estimated the sensitivity at 91-96%, and the specificity at 86%-96%, with the sensitivity for distal occlusions being lower than the sensitivity for proximal occlusions, as expected. However, most stroke centers, including tertiary centers, are reliant on residents and general radiologists, who may be less experienced, and achieve lower sensitivity and specificity, and longer reading times, especially if they are covering additional practices such as a busy trauma center.

In this study, the sensitivity of the RAPID.AI software for proximal occlusions was 88.57% [73.26, 96.8], which is close to the sensitivity of experienced radiologist, however the low specificity translates to a high false positive rate, which may lead, in our experience, to alert fatigue and reduce the overall efficacy of the system.

In the distal occlusion group (m2/m3), the sensitivity was 35.71%. This group is of particular interest given the recent evidence demonstrated that these distal artery occlusions(DAO) are not necessarily “benign” condition. Distal artery occlusions may present with high clinical severity and projected disability. These studies concluded that endovascular therapy for DAO may be safe ^23,24^, hence the importance that the same standards of specificity and sensitivity of proximal LVO will apply on distal occlusions. Given the low sensitivity of radiologist in detecting distal vessel occlusions an AI software can be helpful. However, at the moment sensitivity of 35.71% is likely not sufficient to improve clinical workflow for such distal occlusions.

When reviewing the false positive results, our hypothesis was that poor technical study was the cause, either motion artifact or the timing of contrast injection. However, the results demonstrated that the hypothesis was wrong. Only one study had motion artifacts that were considered moderate, the rest of the studies were without any technical flaw. Most of the false positive alerts resulted in patients with severe chronic stenosis in the ICA or the M1 and were detected as occlusions. Given the fact that the RAPID LVO algorithm abnormality detection is based on drop in the vessel density and comparison to the opposite hemisphere, severe stenosis either in the presumed affected side or the contralateral hemisphere can produce false positive alert. In two cases a subdural hematoma was detected, we assume that the presence of extra axial collection can also effect on the vessel density and produce the false detection alarm.

Another outstanding concern is the long processing time evidenced in our practice - 32.53 min on average in this study and even when excluding 3 cases whose alerts had arrived in a delay, the average time to notification was 18.31 minutes which is longer than was reported in the previous publications for the same software that was 2.86min and much longer compared to published estimates of similar software (e.g., 5:45 minutes for Viz LVO, reported by Shalitin et al.). Anecdotally, the alerts that were excluded arrived hours, or even days, after the scan was performed.

Since time to treatment is a key determinant of patient outcomes, we speculate that long processing times may attenuate the clinical utility of the device.

To the best of our knowledge this is the first big cohort study performed at a big tertiary stroke center, that evaluates the time to notification of RAPID LVO.

Our study has several limitations: First, this is a retrospective design, providing only device performance metrics without measuring the downstream impact on patient outcomes. Second, it is a single site study, evaluating the performance of a single setup. Including both the make and model of the CT scanner, the imaging protocol, as well as the technical setup of the RAPID LVO software, including data transfer and processing setup. These may differ comparing to a different stroke center. A third limitation is the lack of sufficiently large subgroups for posterior circulation occlusions (2 patients), tandem occlusions (1 patient) and isolated ICA occlusions (4 patients).

Further studies including more patients, more sites, emphasis on distal occlusions and different subgroups along with comparison study to other available software could prove insightful and useful.

## Conclusion

In conclusion, at the Montreal neurological hospital (MNH), RAPID LVO has moderate sensitivity, moderate specificity and high time-to-notification performance. Our study data demonstrated in particular moderate overall sensitivity (63%) and low sensitivity for distal occlusions(M2-3), high performance for distal occlusions is of particular importance in light of the emerging evidence on the efficacy of thrombectomy for patients suffering from such distal occlusions. Aside from accuracy, the processing time of the system is a critical determinant of its clinical efficacy, and is likely to affect the clinical utility of the device in setups similar to MNH’s. The disparity between the observed performance and the performance reported in different studies demonstrates the importance of independent, real-world evaluations such as this one. The gap between the performance in this study compared to published records of RAPID AI may be due to differences in imaging hardware, software implementation, connectivity or other technical parameters. A large scale performance study covering a wide range of scanners and sites is required to establish the robustness of performance across a variety of clinical and technical settings.

## Supporting information

Supplemental results Excel

## Data Availability

1. The Imaging are fully available on the MUHC PACS database.
2. The excel spreadsheet is available as supplementary .

## Notes

### Competing Interest Statement

authors #1 and #2 are have received financial support from Viz.ai

### Clinical Trial

The study is retrospective.

### Funding Statement

The study was not funded.

### Author Declarations

On 2021-01-21, delegated review of the research project was provided by members of McGill University Health Centre(MUHC), Research Ethics Board(REB), more precisely its Neurosciences and Psychiatry panel(NEUPSY). After reviewing the documents this research was approved. The Co Chair of the Neurosciences and Psychiatry panel full name is: Brigitte Paquet.

### Summary of Updates

Correction of the conflict of interest.

